# Trends in Gambling-Related Primary Care Encounters in Brazil, 2014–2025

**DOI:** 10.64898/2026.11.09.26367625

**Authors:** Bruno Rezende Souza, João Sérgio da Fonseca Guimarães, Matheus Magno Rosa de Assis, Eduarda Maria Silva de Souza, Luíza Reis Amaral Fonseca, Ana Clara Vagna Pires, Isadora Rodrigues de Carvalho, Kelly Cristina Corrêa dos Anjos, Maria Alice da Costa Nascimento, Maria Clara Silva Barbosa Lima, Maria Clara Diniz Castro, Maria Eduarda Monteiro de Almeida Ferreira, Vitoria Formiga Mansur, André Gustavo Oliveira

## Abstract

The rapid expansion of online gambling and sports betting has raised concerns about gambling-related harm and its implications for public health. However, nationwide evidence regarding temporal trends and patterns of health care utilization associated with gambling remains limited, particularly in low- and middle-income countries. We conducted a nationwide retrospective study using administrative data from Brazil’s Unified Health System from 2014 through 2025. We examined primary health care records associated with pathological gambling (ICD-10, code F63.0) and gambling and betting (code Z72.6). We analyzed temporal trends, seasonal patterns, and geographic and demographic distributions, calculating annual encounter rates per 100,000 inhabitants. Gambling-related primary health care encounters increased substantially during the study period, with a marked acceleration beginning in 2022. By 2025, nationwide rates reached 1.46 encounters per 100,000 inhabitants for pathological gambling and 0.60 per 100,000 for gambling and betting. Increases were observed across all five Brazilian regions, with the highest rates in the South and Southeast. Recorded encounters increased across age groups and both sexes, particularly among adults aged 20 to 39 years. Rates were also elevated among adolescent males and older women. Time-series analyses suggested seasonal variation, with higher indices generally between April and October. Gambling-related encounters in Brazilian primary health care increased markedly from 2014 through 2025, particularly after 2022, with substantial demographic and geographic variation. These findings indicate increasing documentation of gambling-related conditions within a nationwide public health system and underscore the need for improved surveillance, early identification, and access to appropriate mental health care.

## Introduction

Gambling disorder is an increasingly important public health concern, characterized by persistent or recurrent gambling behavior despite substantial personal, financial, and social consequences (World Health Organization (WHO), 2024). The rapid expansion of online gambling platforms and sports betting has substantially increased the accessibility of gambling, raising concerns about its implications for mental health. A recent systematic review and meta-analysis estimated that 46.2% of adults worldwide had participated in gambling during the preceding year, with 1.41% engaging in problematic gambling (Tran et al., 2024). These findings underscore the substantial global exposure to gambling and the need to understand its health consequences, particularly in countries where digital gambling has expanded rapidly.

The clinical burden of gambling disorder extends beyond financial losses and includes considerable psychiatric morbidity, functional impairment, and increased mortality. Gambling disorder is strongly associated with substance use, mood, and anxiety disorders, with a recent meta-analysis estimating that 82.2% of affected individuals had at least one co-occurring psychiatric disorder (Galeazzi et al., 2025). Gambling-related harms have also been associated with suicidal behavior, family disruption, financial insecurity, and socioeconomic inequalities (Wardle et al., 2024). Despite these consequences, gambling disorder remains insufficiently characterized in health care surveillance, particularly in low- and middle-income countries, where information about its clinical burden and patterns of health service utilization is limited.

Brazil provides an important setting for investigating the epidemiology of gambling disorder. The expansion of online betting has raised public health concerns in a country marked by substantial socioeconomic and regional inequalities. A nationally representative survey conducted in 2022–2023 found that 17.6% of individuals aged 14 years or older had participated in gambling during the preceding year, whereas 2.8% were classified as engaging in moderate- or high-risk gambling (Madruga et al., 2026). Although these findings indicate substantial exposure to gambling-related harm, population surveys cannot establish the extent to which these problems are recognized, diagnosed, and treated within the health care system. Brazil’s Unified Health System (Sistema Único de Saúde, SUS) provides publicly funded mental health services through primary care, specialized psychosocial care centers, and hospitals, offering an opportunity to examine the recorded clinical burden of gambling disorder across a large and geographically diverse population (Ministério da Saúde, 2026).

Nationwide health care records can complement population surveys by identifying temporal changes in diagnosed disorders, patterns of service utilization, and demographic and geographic differences in access to care. However, the extent to which gambling disorder is represented in routine health care data, and how this representation has changed over time, remains inadequately understood. Such information is essential for assessing health system responses, identifying potentially underserved populations, and guiding resource allocation and prevention strategies. In this nationwide epidemiological study, we used administrative data from the SUS to examine temporal trends and demographic and geographic patterns in health care encounters recorded with gambling disorder in Brazil.

## Methods

### Data collection

All data used in this study were obtained from the Primary Health Care Information System of the Brazilian Unified Health System (SIAPS/SUS) managed by the Secretariat of Primary Health Care (SAPS). We selected the two most prominent conditions associated with gambling habits classified by the International Classification of Diseases (ICD-10): Pathological gambling (F63.0) and Gambling and betting (Z72.6). We selected the following interest variables: Age group, Sex, Race/ethnicity, Geographical region, and ICD-10 code; and the parameter: number of outpatient visits. Because we could not download data for each analysis as a single dataset, we organized the data into separate datasets: national, regional, and time-series analyses shared a single dataset, whereas age-group, race/ethnicity, and educational attainment analyses each used a separate dataset.

The database comprises consolidated information sent by primary healthcare units, consisting of anonymized clinical assessments documented in primary health care, and should not be interpreted as definitive diagnoses established through specialized psychological or psychiatric evaluation. Data were retrieved from 2014 to 2025 by month/year of the patient visit and reflect the number of unique patients each month to reduce repeated patient visits. Public primary health care served 72% of the Brazilian population in 2024 (https://bvsms.saude.gov.br/bvs/publicacoes/relatorio_anual_gestao_2024.pdf), representing a large share of the population. We do not have access to any individual patient data or information, only aggregate population-level data.

Regional divisions are valid and represent official administrative divisions proposed by the Brazilian Institute of Geography and Statistics (Instituto Brasileiro de Geografia e Estatística–IBGE). We obtained population data from IBGE censuses and estimates throughout the analysis and used them to calculate rates per 100,000.

#### Data Analysis

Time-series analysis was done through the “Forecast” package, analyzing raw outpatient visit numbers, since monthly population numbers were not available. To calculate rates per 100,000 patient visits, we summed monthly visits within each year to accurately adjust for population, since population data are available by year. Race/Ethnic data were normalized by the gender-specific national population, because specific racial population counts/estimates were not available for all analyzed years. Data were analyzed using R (version 4.4.2) and RStudio (2024.04.2 Build 764), and data visualization was performed through the “geobr” and “ggplot2” packages.

For the Brazilian regional maps and age-dependent analysis, we chose 2014, 2018, 2022, and 2025 because they mark key years in Brazil’s gambling legislation. 2014 marks the first year of control; 2018 is when fixed-odds betting was legalized (“LEI N° 13.756,” 2018); 2022 was the deadline for betting companies to register under the law (“LEI N° 14.790,” 2023); and 2025 is the latest available year with complete data under the 2025 sports-betting regulatory framework (“PORTARIA SPA/MF N° 817,” 2025).

Because SIAPS serves 72% of the population and we corrected data using official government counts and estimates, we avoided statistical testing when comparing the impact of conditions across regions, sexes, races, and age groups.

## Results

### Time-Series and geographical analysis of gambling-related conditions

Outpatient visits for Pathological Gambling (F63.0) showed a low trend at the start of the analysis period in 2014, but increased sharply in the latter half of 2022 and remained high through 2025 (Fig. 1A). Visits due to Gambling and Betting (Z72.6) also showed a similar dynamic, with the initial increase during 2022, albeit with a lower number of visits in the same period (Fig. 1B). Interestingly, analysis of the seasonal index component for Pathological Gambling reveals a growing pattern from April to October, with a positive index from July to October, and a decrease from November to March (Fig. 1C). Gambling and betting showed a less stable but still noticeable growth throughout the months, with the seasonality index increasing from March to October and declining from November to February (Fig 1D).

**Figure 1:**
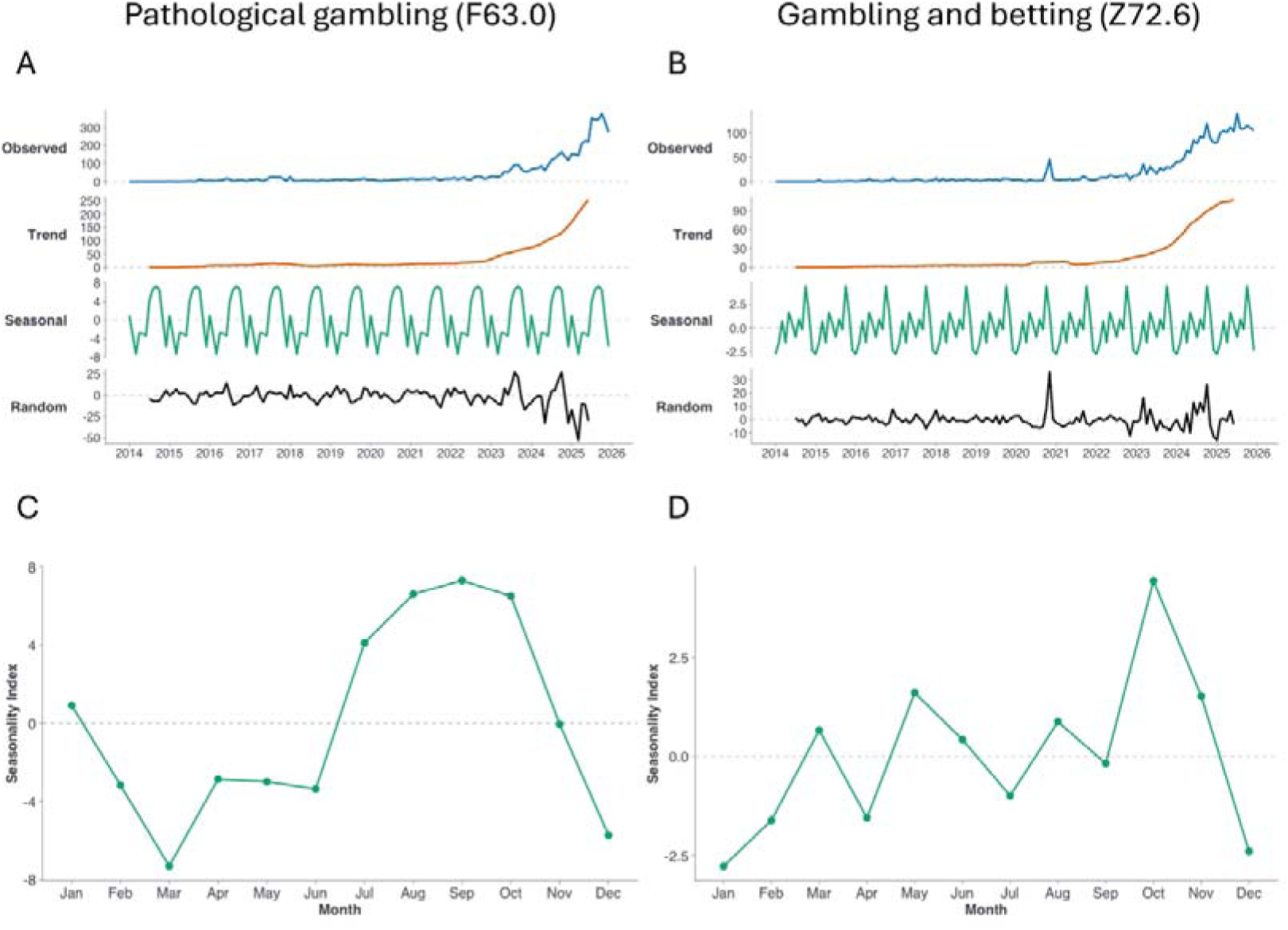
Dynamics of Outpatient visits due to Gambling disorders in Brazil. (A-B) Dynamics of Outpatient visits for Pathological Gambling (F63.0) and Gambling and Betting (Z72.6), respectively. Both conditions showed similar trends: a low point in 2014, followed by an increase in 2022, and continued through 2025. (C-D) Monthly seasonality index for Pathological Gambling (F63.0) and Gambling and Betting (Z72.6), respectively. Note that the scales between panels A and B are different.

Geographical data show a shared pattern between F63.0 and Z72.6 for outpatient visits per 100,000 people, despite scale differences: both conditions grew across all Brazilian regions, with the South and Southeast showing the highest incidence by 2025. Regional rates varied from 0.79 to 1.84 visits per 100,000 for F63.0 (Fig. 2A) and from 0.37 to 0.87 visits for Z72.6 in 2025 (Fig. 2B). Rates per 100,000 reached similarly high levels in the nationwide context, showing a reduced value during the initial years and reaching, in 2025, 1.46 and 0.60 outpatient visits per 100,000 people for F63.0 and Z72.6, respectively (Fig 2C). When comparing the nationwide and regional subsets for both conditions, we found similar regional trends that also reflected the national dynamics for F63.0 and Z72.6

**Figure 2:**
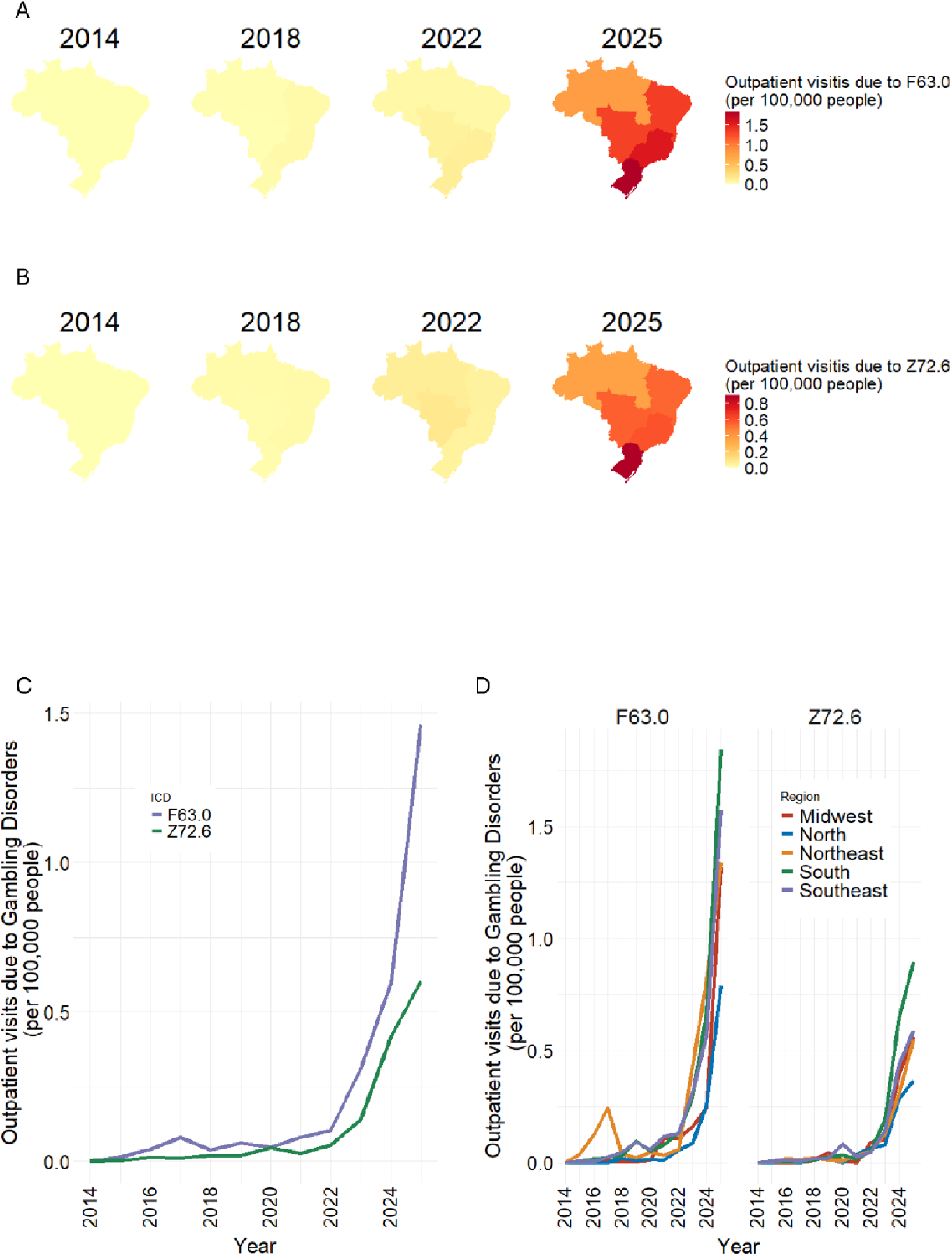
Outpatient visits due to Gambling disorders in Brazil and its regions. (A-B) Outpatient visits for Pathological Gambling (F63.0) and Gambling and Betting (Z72.6), respectively, per 100,000 people in 2014, 2018, 2022, and 2025 across all Brazilian regions. C) Outpatient visits due to Gambling disorders per 100,000 people from 2014 to 2025 in Brazil. D) Outpatient visits due to Gambling disorders per 100,000 people from 2014 to 2025 across all Brazilian regions. Note that the color-gradient scales between panels A and B are different.

### Sex, Racial/Ethnic and Age-Specific burden of gambling-related conditions

We analyzed how socio-demographic parameters affected outpatient visit rates across the population. For age- and sex-dependent variables, no outpatient visits were registered in 2014 (Fig. 3). By 2018, men aged 15-19 (F63.0: 0.043 and Z72.6: 0.018) and 50-59 (F63.0: 0.043 and Z72.6: 0.017) showed a slight increase, mostly for F63.0. For women, the increase was higher among ages 50-59 (F63.0: 0.039 and Z72.6: 0.004), 60-69 (F63.0: 0.045 and Z72.6: 0.032), and 70-79 (F63.0: 0.036 and Z72.6: 0.024), mostly for F63.0 (Fig. 3). By 2022, all ages in both sexes increased. In men, the largest increase was among those aged 20-29 years, for both F63.0 and Z72.6 (F63.0: 0.149 and Z72.6: 0.091) (Fig. 3). For women, the largest increase was among those aged 70-79, mostly for F63.0 (F63.0: 0.062 and Z72.6: 0.010) (Fig. 3). By 2025, rates for both F63.0 and Z72.6 increased sharply for both sexes and all ages. Notably, men show an earlier onset, with ages 10-14 (F63.0: 0.418 and Z72.6: 0.068) and 15-19 (F63.0: 0.908 and Z72.6: 0.488) already showing increased outpatient visits, while women remain relatively low at 10-14 (F63.0: 0.418 and Z72.6: 0.021) and 15-19 (F63.0: 0.156 and Z72.6: 0.054) (Fig. 3). From ages 20-29 (Men-F63.0: 1.749 and Z72.6: 0.867; Women-F63.0: 1.286 and Z72.6: 0.632), 30-39 (Men-F63.0: 1.420 and Z72.6: 0.691; Women-F63.0: 1.558 and Z72.6: 0.719), and 40-49 (Men-F63.0: 0.673 and Z72.6: 0.235; Women-F63.0: 0.798 and Z72.6: 0.247), both sexes show a substantial increase, representing the largest fraction of outpatient visits (Fig. 3). Ages 50 onward show a relative reduction compared with previous age groups, but higher rates in women than in men (Fig. 3).

**Figure 3:**
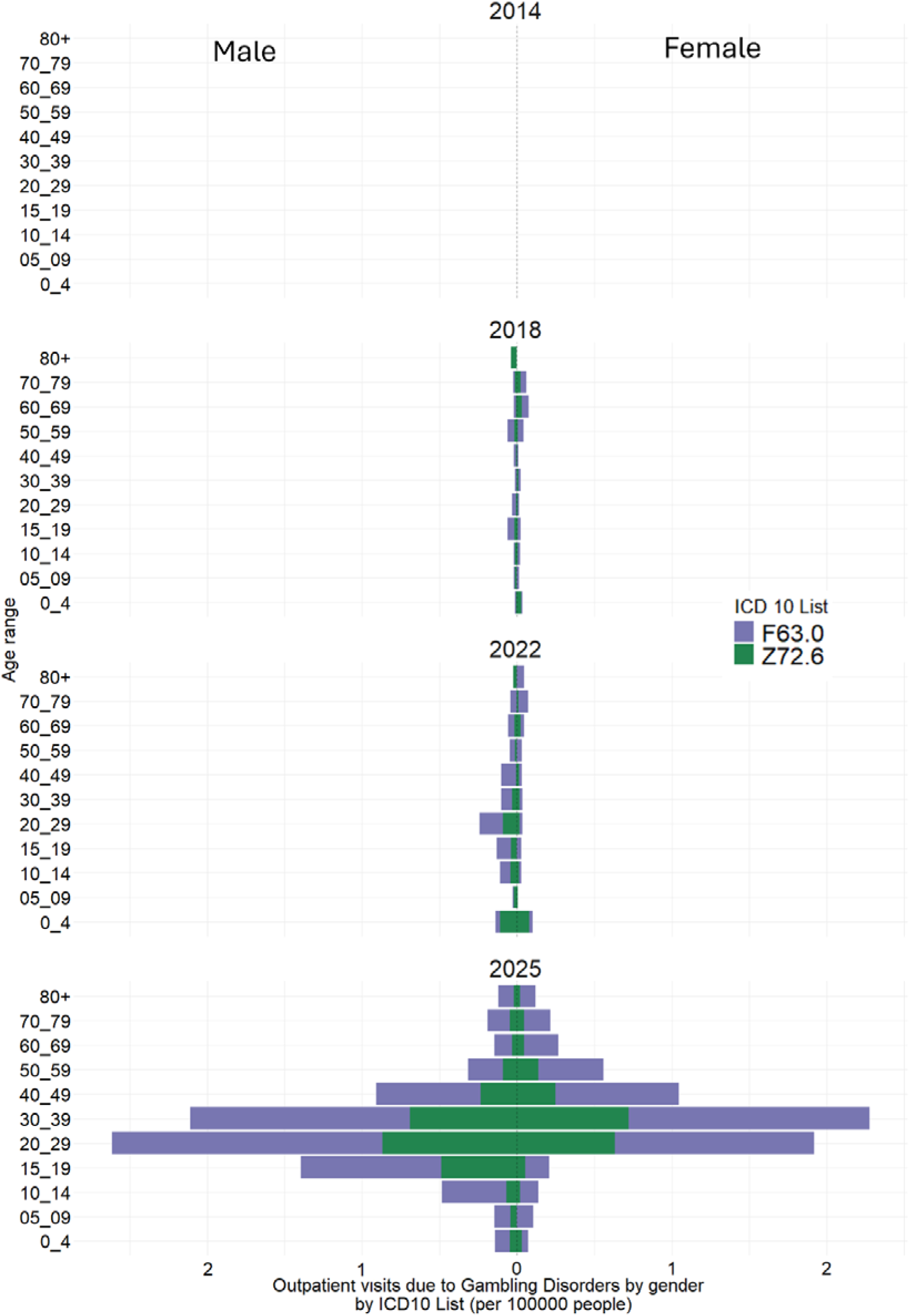
Outpatient visits due to Gambling disorders by age, sex, and ICD10 classification per 100,000 people in 2014, 2018, 2022, and 2025 in Brazil.

When analyzing outpatient visits in the racial/ethnic subset, we identified similar profiles between black and white populations regarding Pathological gambling (F63.0). We didn’t observe gender differences within either race. In 2025, black men and women reached 0.33 and 0.36 outpatient visits per 100,000 people, respectively, while white men and women reached 0.33 and 0.23 visits per 100,000 people, respectively. Asian and Native-American populations had markedly lower rates, with Asian men and women at 0.02 and 0.01, respectively, while Native-American men had 0 visits in 2025 and Native-American women had 0.001 (Fig. 4A). Gambling and betting (Z72.6) show a similar trend, with the white and black populations having similarly high levels in 2025, with black men and women reaching 0.16 and 0.14 outpatient visits per 100,000 people, respectively, and white men and women reaching 0.14 and 0.11 visits. Asian populations showed 0.007 and 0.005 outpatient visits per 100,000, and the Native American population recorded no visits during the entire period (Fig. 4B).

**Figure 4:**
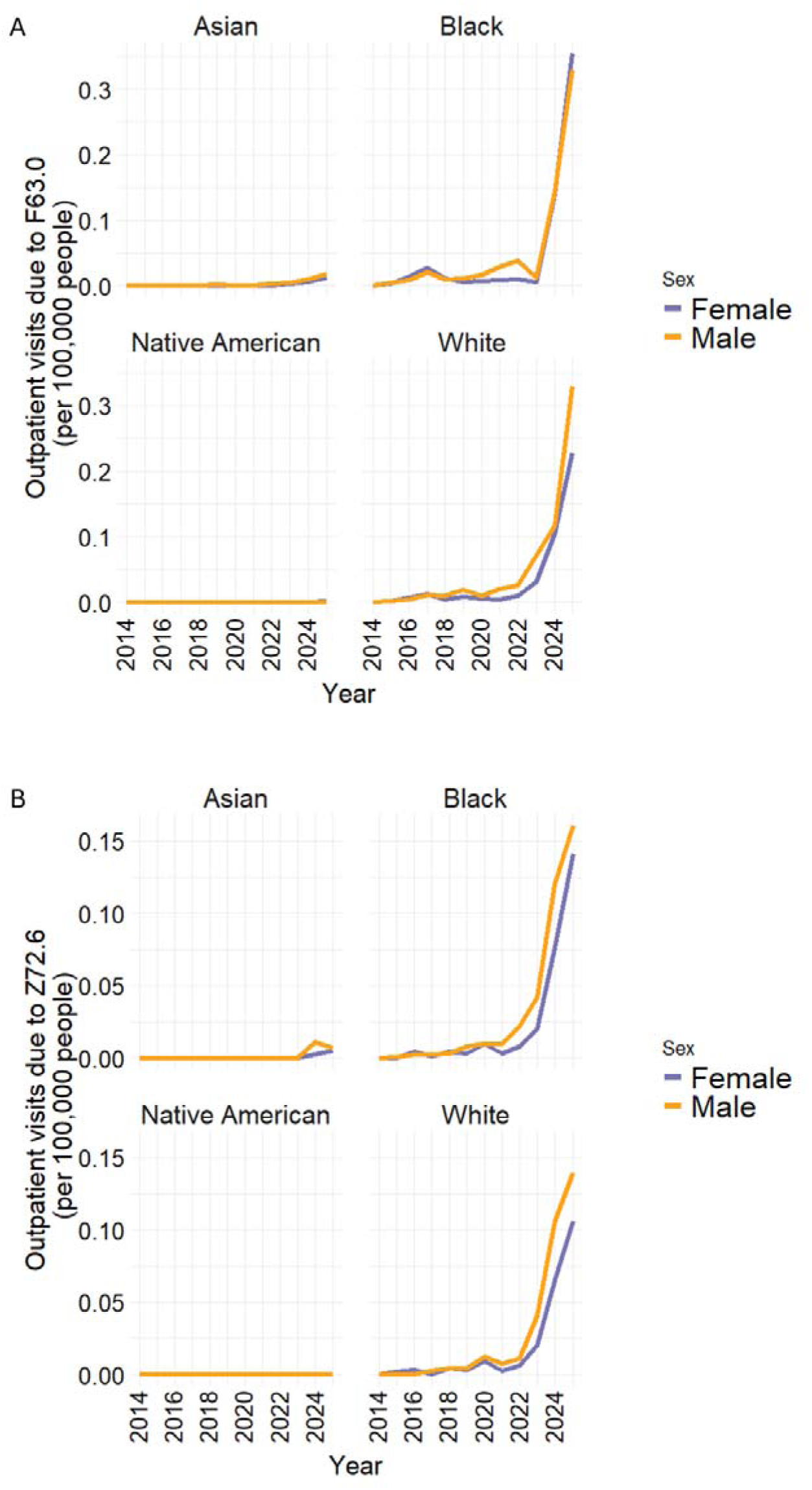
Outpatient visits due to Gambling disorders by race/ethnicity and sex. A) Outpatient visits for Pathological Gambling (F63.0) by race/ethnicity and sex per 100,000 people from 2014 to 2025 in Brazil. B) Outpatient visits for Gambling and Betting (Z72.6) by race/ethnicity and sex per 100,000 people from 2014 to 2025 in Brazil.

## Discussion

In this nationwide retrospective study of gambling-related primary health care encounters in Brazil, we identified an increase in records associated with pathological gambling (ICD-10 F63.0) and gambling and betting (ICD-10 Z72.6) between 2014 and 2025, particularly after 2022. By 2025, national rates had reached 1.46 and 0.60 recorded encounters per 100,000 inhabitants, respectively. Increases were observed across all five geographic regions and among both sexes, with particularly prominent changes among young adults. These findings are consistent with growing concerns about gambling-related harm worldwide. A systematic review and meta-analysis estimated that 46.2% of adults had gambled during the preceding year and that 1.41% experienced problematic gambling (Tran et al., 2024). In Brazil, the Third National Alcohol and Drugs Survey (LENAD III) found that 17.6% of individuals aged 14 years or older had gambled during the preceding year, with 2.8% classified as moderate- or high-risk gamblers (Madruga et al., 2026). Although these population estimates are not directly comparable with clinical encounters, our findings document an increasing representation of gambling-related conditions in Brazilian primary health care.

The marked increase in gambling-related encounters occurred during substantial changes in the availability, accessibility, and regulation of gambling in Brazil. The expansion of online gambling has transformed participation by enabling continuous access through mobile devices, reducing geographic barriers, and increasing opportunities for rapid, repeated betting (Ghelfi et al., 2023; Wardle et al., 2024). In Brazil, fixed-odds sports betting was legalized in 2018 (“LEI N° 13.756,” 2018), followed by broader regulation of online gambling in 2023 (“LEI N° 14.790,” 2023) and implementation of federal authorization requirements in 2025 (“PORTARIA SPA/MF N° 817,” 2025). The temporal increase identified in our study is particularly noteworthy because it became more pronounced in 2022, before the implementation of the most recent regulatory framework. The LENAD III survey identified online gambling as a particularly strong correlate of gambling-related risk (Tran et al., 2024), suggesting that changes in gambling accessibility may have important public health implications. Changes in awareness, health-seeking behavior, diagnostic practices, and the completeness of administrative records may also have contributed to the observed trends.

The increase in gambling-related primary health care encounters is particularly relevant when considered alongside the broader trajectory of psychiatric health care in Brazil. In a recent nationwide study of the Brazilian Unified Health System, Resende et al. reported that psychiatric hospitalization rates declined by approximately 52% between 2008 and 2022 (Resende et al., 2026). In contrast, this study identified an increase in gambling-related primary-care records, beginning particularly in the latter part of 2022 and continuing through 2025. Although the two studies evaluated different levels of care and their observation periods overlapped only partially, these findings suggest potentially divergent patterns of mental health service utilization. Reductions in psychiatric hospitalizations should not necessarily be interpreted as evidence of declining psychiatric morbidity, particularly for disorders that are increasingly recognized or managed in outpatient settings. The growing documentation of gambling-related conditions may therefore represent an additional challenge for Brazil’s community-based mental health services.

Despite the increase observed during the study period, the absolute rates of gambling-related primary-care encounters remained low relative to population-based estimates of gambling-related risk. This discrepancy is consistent with previous evidence showing that many individuals experiencing gambling-related problems do not receive professional treatment. A systematic review found that gambling disorder is rarely assessed systematically in primary care and that general practitioners frequently report insufficient knowledge, confidence, or training in its management (Tatar et al., 2025). Previous research has identified stigma, limited awareness of treatment services, and reluctance to disclose gambling problems as barriers to help-seeking (Gainsbury et al., 2014). An additional consideration is that gambling-related harms may not be recorded under gambling-specific diagnostic codes when individuals present with associated psychiatric or physical health complaints. A recent meta-analysis estimated that 82.2% of individuals with gambling disorder had at least one co-occurring psychiatric disorder, particularly substance use, mood, and anxiety disorders (Galeazzi et al., 2025). Gambling-related problems have also been associated with suicidal ideation, suicide attempts, and suicide mortality (Kristensen et al., 2024). Consequently, the relatively low rates identified in the present study may reflect incomplete recognition and documentation of gambling-related problems within routine primary care rather than a correspondingly low population burden. Importantly, the two ICD-10 categories examined here represent different clinical constructs: F63.0 refers to pathological gambling, whereas Z72.6 records gambling and betting without necessarily indicating a psychiatric disorder (World Health Organization (WHO), 2019).

The demographic distribution of gambling-related encounters provides additional insights into populations potentially requiring targeted prevention and treatment. By 2025, most age groups in both sexes showed increases, with particularly high rates among adults aged 20 to 39 years. Among males, elevated rates were also recorded during adolescence, whereas women showed relatively higher rates in some older age groups. These findings are consistent in part with recent Brazilian survey evidence. In a nationally representative study of online betting participation, Magini et al. found that adults aged 18 to 34 years had substantially higher odds of online betting than adults aged 60 years or older, and that men had higher odds of participation than women (Magini et al., 2026). Evidence from a systematic review of gambling during childhood, adolescence, and emerging adulthood also identified increased vulnerability among young people, particularly males, and associations between novel gambling activities and problematic gambling (Richard and King, 2023). However, patterns of gambling participation should not be equated with patterns of gambling-related health care utilization. Differences in clinical recognition, help-seeking behavior, and symptom severity may contribute to the age- and sex-specific distributions we identified. The presence of gambling-related clinical records among adolescents is particularly concerning and supports the importance of early identification and prevention.

Geographic analyses revealed increasing gambling-related encounters across all Brazilian regions, with the highest recorded rates in the South and Southeast by 2025. These differences may reflect variations in gambling accessibility, socioeconomic conditions, digital connectivity, health care infrastructure, and the recognition of gambling-related conditions. A systematic review examining inequalities in gambling-related harms found that socioeconomic status, age, education, ethnicity, and gambling behavior were associated with differences in the distribution of harm (Raybould et al., 2021). Moreover, gambling-related harm extends beyond clinical diagnoses and may include financial insecurity, family conflict, psychological distress, occupational impairment, and social disadvantage (Langham et al., 2015). Therefore, geographic differences in recorded encounters may reflect both variations in gambling-related morbidity and inequalities in access to health care. The lower rates documented in some regions cannot be interpreted as evidence of a lower underlying burden without accounting for differences in service availability and clinical ascertainment. Similarly, interpret the racial and ethnic distributions observed in our study cautiously because appropriate race-specific population denominators were not available throughout the study period. These limitations reinforce the need to strengthen epidemiological surveillance among historically underserved populations.

The time-series analyses also suggested seasonal variation in gambling-related encounters, with higher seasonal indices generally observed between April and October. This pattern may partly reflect the Brazilian football calendar, which provides sustained opportunities for sports betting throughout much of the year. State football championships traditionally begin in January, while the Brazilian National Championship (Campeonato Brasileiro Série A) generally begins in March or April and runs through the final months of the year. In 2025, for example, state championships began in January, and the national championship started on March 29 ((CBF), 2024). The shift from regional competitions to a longer national football season could contribute to sustained betting exposure during the middle and latter portions of the year. Nevertheless, this interpretation remains speculative. The available data do not identify individual gambling activities, betting frequency, or the temporal relationship between gambling exposure and clinical encounters. Seasonal variation in financial circumstances, help-seeking behavior, and health service utilization may also contribute to the observed patterns. Studies integrating gambling activity, sporting calendars, and clinical data are needed to investigate these associations.

The increasing documentation of gambling-related conditions has important implications for mental health policy and primary-care practice in Brazil. Gambling disorder is associated with substantial psychiatric morbidity (Galeazzi et al., 2025), and suicidal behavior represents an important clinical concern among affected individuals (Kristensen et al., 2024). International clinical guidelines recommend the identification of gambling-related harm, assessment of psychiatric comorbidities and suicide risk, and the establishment of referral pathways to specialized treatment when appropriate ((NICE), 2025). Primary health care may represent an important setting for these interventions, particularly because patients experiencing gambling-related problems may initially present with anxiety, depression, substance use, or financial and interpersonal difficulties. The integration of gambling-related assessment into routine mental health care, professional training, and community-based referral services could improve clinical recognition. At the population level, prevention strategies should also address environmental and commercial determinants of gambling-related harm, including advertising, accessibility, and product design (Wardle et al., 2024). These measures may be especially relevant in Brazil, where the rapid expansion of digital gambling has occurred alongside persistent socioeconomic and geographic inequalities.

Our study has several limitations. First, we used aggregated administrative data that did not include individual clinical histories, validated diagnostic assessments, or information regarding gambling severity and duration. Second, the records represent documented primary-care encounters rather than all individuals experiencing gambling-related harm, and therefore cannot establish the incidence or prevalence of gambling disorder. Third, although the data identify unique individuals within each month, the same person may appear in multiple months, preventing the estimation of annual unique-patient counts. Fourth, the lack of individual-level information on gambling modalities, socioeconomic circumstances, psychiatric comorbidities, and treatment limited the interpretation of demographic and geographic differences. Finally, the descriptive design does not permit causal attribution of changes in health care utilization to gambling market expansion, regulatory developments, or specific sporting events.

In conclusion, gambling-related primary health care encounters increased substantially in Brazil between 2014 and 2025, particularly after 2022, with considerable demographic, geographic, and seasonal heterogeneity. This increase occurred against a historical background of declining psychiatric hospitalizations nationwide, highlighting the evolving demands placed on Brazil’s mental health system. Although the findings cannot establish changes in the population prevalence of gambling disorder, they document increasing clinical recognition or recording of gambling-related conditions within the public primary-care system. Strengthening surveillance, integrating gambling-related assessment into mental health services, and expanding equitable access to prevention and treatment may help address the emerging public health challenges associated with gambling.

## Data Availability

All data produced are available online at: https://siaps.saude.gov.br/publico/praticas-assistenciais

## Acknowledgments

To the young scientists at the NeuroDEv and Physiological Circuits laboratories, thank you for your daily support. To our colleagues, we appreciate our scientific discussions and your assistance during times of urgency. And to the often “invisible” yet indispensable members of our university community, including technicians, cleaning personnel, the security team, restaurant staff, and administrative professionals. Your contributions are fundamental to our research endeavor.

